# The evaluation of the clinical effect of primary insomnia treated with the manual acupuncture therapy: a study protocol for a parallel, randomized controlled trial

**DOI:** 10.1101/2024.04.16.24305887

**Authors:** Kean Zhu, Chuanlong Zhou, Quanai Zhang, Dexiong Han, Weiji Chen, Wenfang Wang, Bin Wu, Xianming Lin, Yingjun Liu

**Author notes:** Corresponding author (LYJ). These authors contributed equally to this work.

## Abstract

**Background:** Clinically, primary insomnia (PI) manifests as difficulty falling asleep, difficulty maintaining sleep, or early waking up. The occurrence of PI is closely related to emotional disorders, social stress and environmental stress. According to current studies, acupuncture offers significant advantages in treating PI. The manual acupuncture has been used for the treatment of insomnia for decades. Different acupuncture strategies, including electroacupuncture, manual acupuncture, and multiple acupuncture, have been reported to treat insomnia. However, there are few standardized clinical reports to investigate the effect of the manual acupuncture and its mechanism.

**Methods:** This study was a parallel, randomized controlled trial designed to investigate the clinical efficacy of the manual acupuncture in treating PI, and to clarify the mechanism. The study included a 4-week treatment period and a 4-week follow-up period. All eligible 74 patients diagnosed with PI will be evaluated using scales before treatment, and then be randomly assigned to the acupuncture group and the placebo acupuncture group at a ratio of 1:1. Both groups choose the same acupoints. The patients will be evaluated at 4 weeks before treatment, after 4 weeks of treatment and 4 weeks after treatment respectively. The patients in the acupuncture group were treated with manual acupuncture and placebo acupuncture group received the tube of Park sham devices. All 12 sessions of treatment will last over 4 weeks (3 sessions per week). The main outcome measure will be the change in Pittsburgh sleep quality index before and after treatment, and the secondary outcome measures include Chalder-14 fatigue scale, Epworth sleepiness scale, self-rating depression scale, self-rating anxiety scale, polysomnography and heart rate variability.

**Discussion:** This study will evaluate the efficacy and safety of the manual acupuncture, and the results will provide a scientific theoretical basis for the clinical promotion.

**Trail registration:** ClinicalTrails.gov Identifier: NCT05830887. Registered 23 March 2023.

## 1. Background

According to the International Classification of Sleep Disorders Third Edition (ICSD-3), primary insomnia (PI) is defined as the phenomenon of difficulty initiating or maintaining sleep under adequate sleep opportunities and suitable sleep environments, accompanied by impaired daytime energy or work performance, but without any secondary factors. Epidemiological surveys have shown that insomnia occur in a high proportion of the adult population, between 35 and 50%[1]. Chronic insomnia is usually associated with anxiety, depression, daytime fatigue, and impaired social/occupational functioning[2–4]. Studies have shown that about 30% of PI patients have depressive symptoms and about 20% have other psychiatric symptoms[5]. At present, drug treatment and non-drug treatment can be included for PI[6]. Drug therapy usually uses sedatives and hypnotics, anti-anxiety and depression drugs, antihistamines, and melatonin et al[7]. However, they are limited by their abundant side effects, such as drug addiction, dizziness, fatigue, increased periodic limb activity during sleep, and aggravation of symptoms after irregular drug withdrawal[8]. As one of non-drug treatments, cognitive behavioral therapy for sleep (CBTi) requires educated patients. The efficacy of CBTi will be obviously reduced if patients cannot understand or correctly implement the sleep behavior correction content of CBTi.

As a complementary and alternative therapy, acupuncture is based on the unique meridian theory and dialectical treatment system of traditional Chinese medicine. PI can be treated with acupuncture, which has many benefits, including lower costs, better effectiveness, and no toxic side effects. As a kind of acupuncture therapy, the manual acupuncture for insomnia advocates “focusing on acupuncture for calming down”. Modern studies have shown that the manual acupuncture can increase release of serotonin in serum, change temporal lobe signals, improve the oxygen saturation of brain in the visual cortex, improve the blood oxygen that brought to the brain, and thus improve the sleep quality[9,10].

After years of clinical promotion, the manual acupuncture is well accepted by patients. The in-depth study of its clinical efficacy could help further promote and apply which to treat PI. However, there is no reported standardized clinical research yet.

## 2. Methods

### 2.1 Study design

This is a parallel, randomized controlled trial (RCT) to explore the clinical efficacy and the mechanism of the manual acupuncture. All eligible subjects (n= 74) will be assessed by the Chalder-14 fatigue scale, Epworth sleepiness scale, self-rating depression scale (SDS), and self-rating anxiety scale (SAS) before treatment, and then randomly assigned to the treatment group (n=37) or the control group (n=37) at a ratio of 1:1. The treatment will be given 3 times a week for 4 weeks. The assessments of the scales, polysomnography (PSG), and heart rate variability (HRV) will be conducted again after four weeks of treatment and once more after a one-month follow-up.

### 2.2 Participants

This RCT will be conducted at the Third Affiliated Hospital of Zhejiang Chinese Medical University. Subjects will be recruited through advertisements on the hospital’s electronic social platform, posters distributed in hospital public areas and voluntary speech with details and contact information for this study. Participants were recruited between April 1, 2023, and January 31, 2025. Subjects are initially screened by contacting an acupuncturist and potential eligible participants will be registered to make an appointment with the designated acupuncturist to check the inclusion and exclusion criteria. All eligible patients were required to sign a written informed consent prior to randomization. The inclusion and exclusion criteria are as follows:

#### 2.2.1 Inclusion criteria

The patients will be included if they meet all of the following criteria:

1. Meets the diagnostic criteria for PI[11];
2. The participants ranged in age from 18 to 70 years old;
3. PSQI score > 7;
4. No communication and cognitive dysfunction;
5. No use or discontinuation of anti-anxiety and other psychotropic drugs within one month;
6. No major physical diseases;
7. Continue to accept the research content and complete the assessment of various scales;
8. Signed the informed consent form before starting the study.

#### 2.2.2 Exclusion criteria

1. Those who did not meet the inclusion criteria;
2. Patients suffering from severe mental disorders, severe head trauma history and accompanied by significant consciousness disturbance;
3. Patients with severe hepatic and renal dysfunction and bleeding tendency;
4. Alcoholism (liquor ≥100ml/ day), smoking (≥15 cigarettes/day), drug abuse or taking psychotropic drugs;
5. Suffering from other sleep disorders, such as sleep apnea-hypopnea syndrome, narcolepsy, rapid eye movement and sleep behavior disorder;
6. Pregnant or lactating;
7. Patients with other major diseases and poor control;
8. Others are unwilling to sign informed consent.

### 2.3 Withdrawal criteria

1. Patients with adverse reactions and intolerance after treatment;
2. Unable or unwilling to continue treatment due to other sudden diseases during the study;
3. Patients whose symptoms deteriorated during treatment, and patients who did not want to continue treatment;
4. Self-withdrawal from the investigator;
5. Failing to complete the treatment within the specified time and unable to observe the indicators;
6. Patients who completed the treatment but failed to follow-up and could not observe the final indicators.

Note: Dropouts are defined as any of the above.

### 2.4 Ethics approval and consent to participate

The clinical protocol and written informed consent were approved by the Medical Ethics Association of the Third Affiliated Hospital of Zhejiang Chinese Medical University (NO. ZSLL-KY-2023-012-01). The study complies with the ethical principles of the Declaration of Helsinki. All the participants gave informed consent to participate in study before taking part.

### 2.5 Randomization

The SPSS16.0 software will be used to generate 74 random numbers. The numbers will be equally distributed into two groups according to a ratio of 1:1, namely 37 digits in the acupuncture group and 37 digits in the placebo acupuncture group. The generated random numbers and grouping information will be transferred to a sealed envelope and kept by the random allocation personnel. When patients undergo multiple screenings of diagnostic criteria, inclusion criteria and exclusion criteria, baseline assessment, “informed consent” is signed, and finally randomized by those responsible for randomization.

### 2.6 Blinding

Participants will be randomly assigned to the acupuncture group and the placebo acupuncture group at a ratio of 1:1. The efficacy was evaluated blindly, and the scales were summarized and scored by the researchers who were unaware of the grouping. Blinded statistics were used at the data summary stage, and research results were statistically analyzed by statisticians unaware of the grouping. In the study, the treatment implementers, the efficacy evaluators, and the data statisticians were processed independently.

### 2.7 Interventions

According to acupuncture theory, the interventions of this research were developed by consensus of acupuncture experts. To ensure the homogeneity of treatment, all acupuncturists participating in this study must be attending doctor or above and will receive special training to fully understand the therapies and operate as the same as the presented acupuncture methods. Both groups choose the same acupoints, the acupuncture manipulations are supported by the Park devices. Patients in both groups were treated in separate rooms. The specific device is shown in figure 1[12]. All 12 sessions will be administered over a 4-week period (3 sessions per week), followed by 4 weeks after treatment. The study plan is shown in table 1.

**Figure 1.**
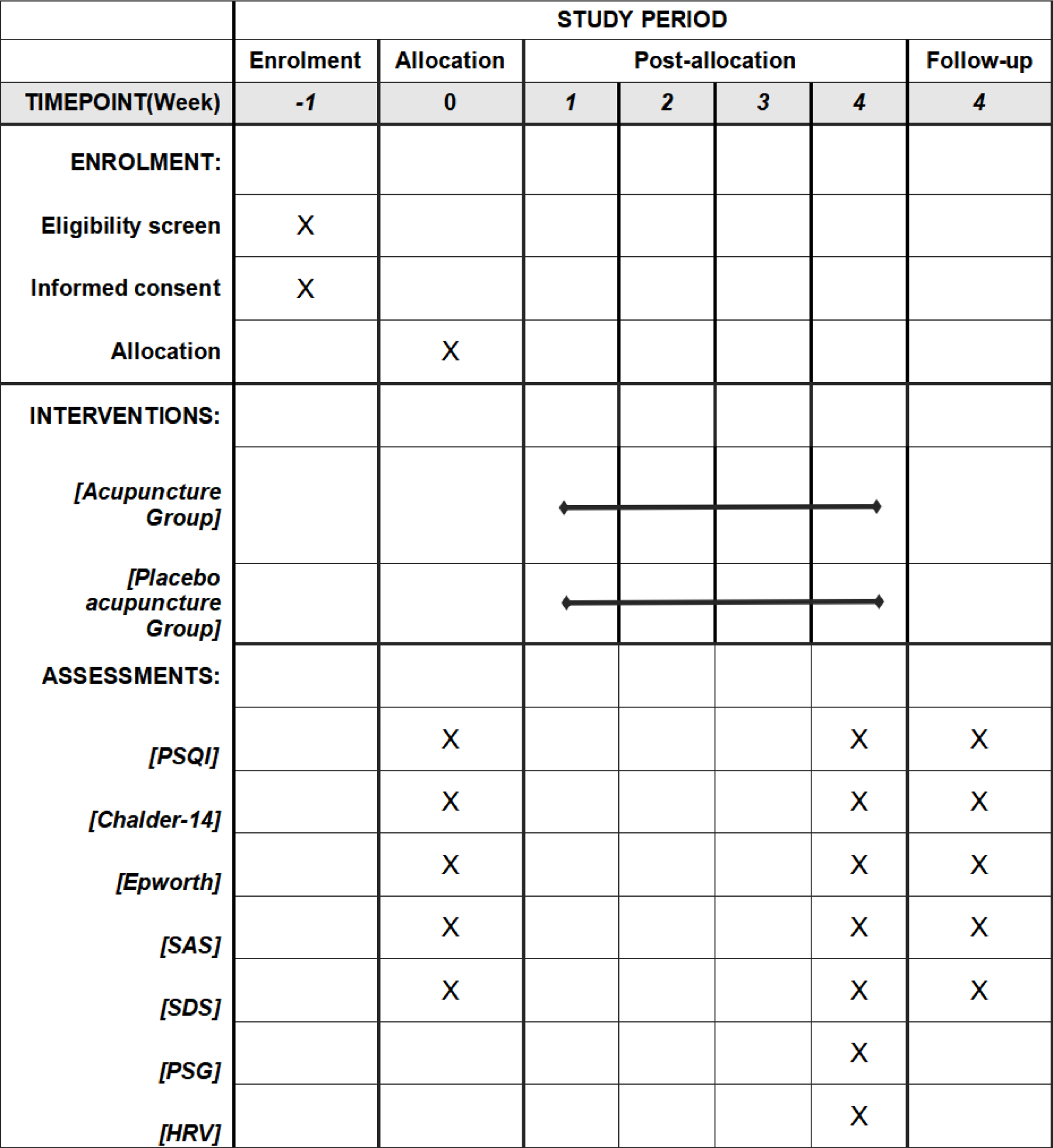
Schedule of enrollment, interventions and assessments.

**Table 1.**
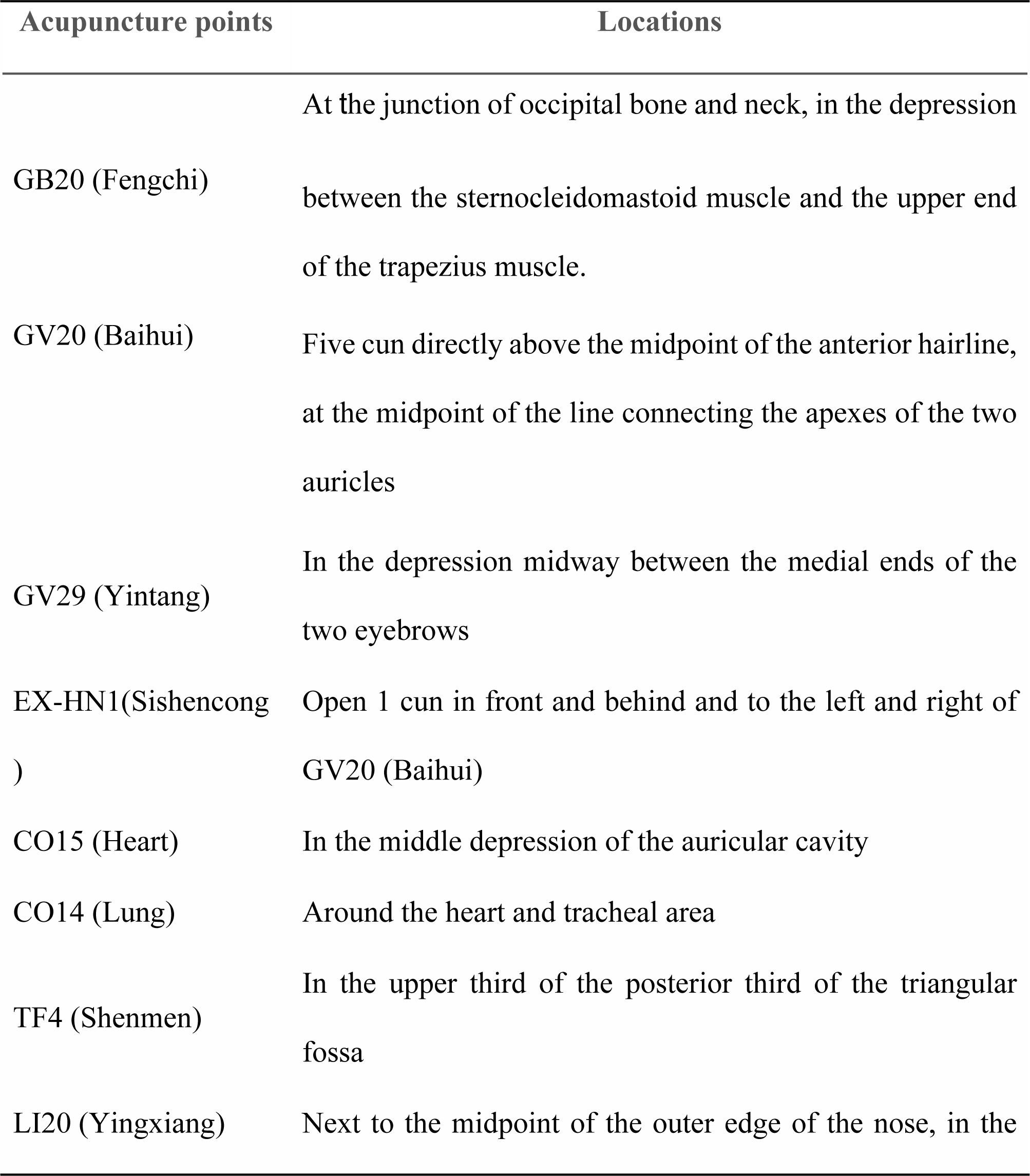
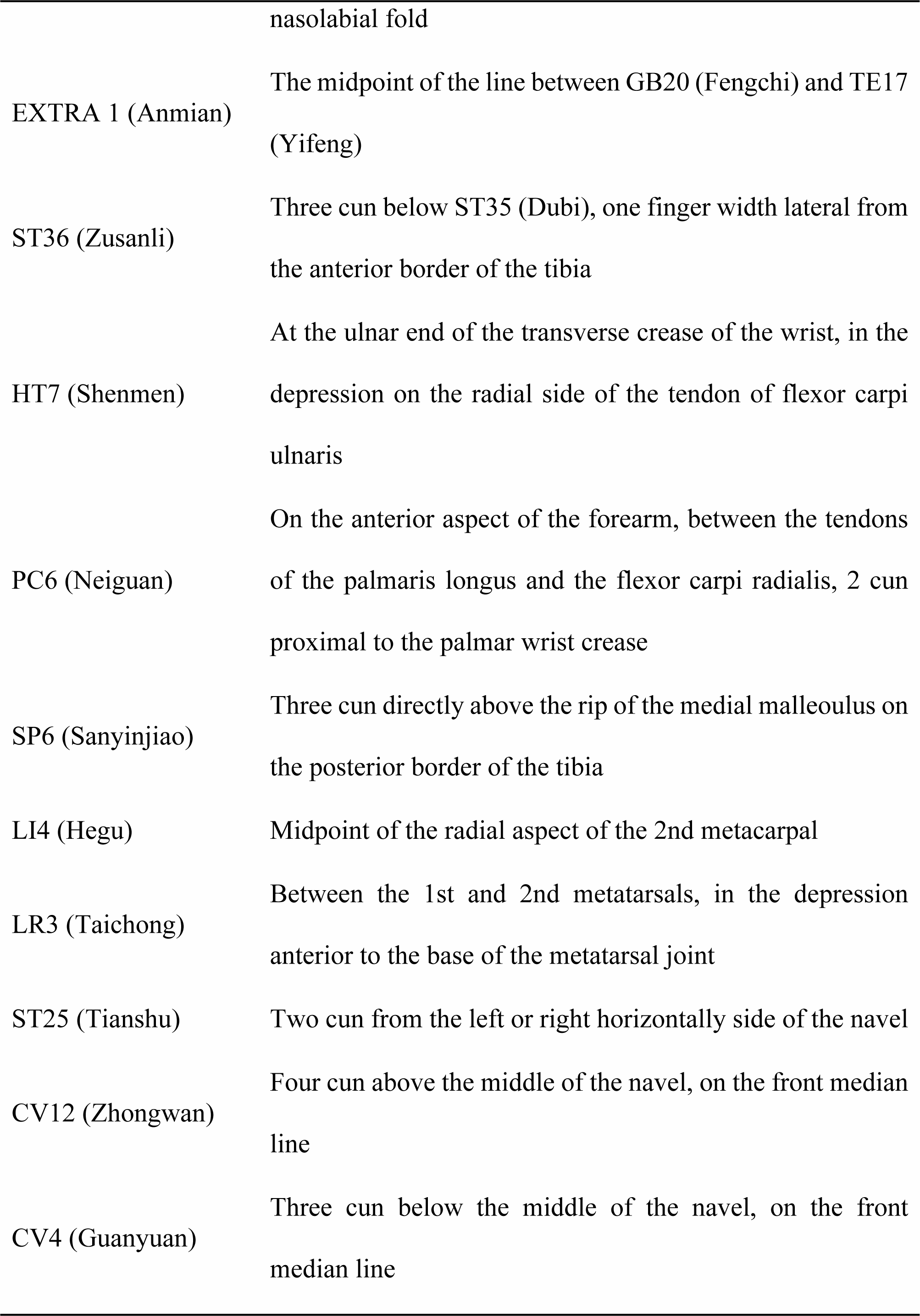
Framework of the acupuncture point prescription.

| Acupuncture points | Locations |
| --- | --- |
| GB20 (Fengchi) | At the junction of occipital bone and neck, in the depression between the sternocleidomastoid muscle and the upper end of the trapezius muscle. |
| GV20 (Baihui) | Five cun directly above the midpoint of the anterior hairline, at the midpoint of the line connecting the apexes of the two auricles |
| GV29 (Yintang) | In the depression midway between the medial ends of the two eyebrows |
| EX-HN1(Sishencong ) | Open 1 cun in front and behind and to the left and right of GV20 (Baihui) |
| CO15 (Heart) | In the middle depression of the auricular cavity |
| CO14 (Lung) | Around the heart and tracheal area |
| TF4 (Shenmen) | In the upper third of the posterior third of the triangular fossa |
| LI20 (Yingxiang) | Next to the midpoint of the outer edge of the nose, in the |
|  | nasolabial fold |
| EXTRA 1 (Anmian) | The midpoint of the line between GB20 (Fengchi) and TE17 (Yifeng) |
| ST36 (Zusanli) | Three cun below ST35 (Dubi), one finger width lateral from the anterior border of the tibia |
| HT7 (Shenmen) | At the ulnar end of the transverse crease of the wrist, in the depression on the radial side of the tendon of flexor carpi ulnaris |
| PC6 (Neiguan) | On the anterior aspect of the forearm, between the tendons of the palmaris longus and the flexor carpi radialis, 2 cun proximal to the palmar wrist crease |
| SP6 (Sanyinjiao) | Three cun directly above the tip of the medial malleolus on the posterior border of the tibia |
| LI4 (Hegu) | Midpoint of the radial aspect of the 2nd metacarpal |
| LR3 (Taichong) | Between the 1st and 2nd metatarsals, in the depression anterior to the base of the metatarsal joint |
| ST25 (Tianshu) | Two cun from the left or right horizontally side of the navel |
| CV12 (Zhongwan) | Four cun above the middle of the navel, on the front median line |
| CV4 (Guanyuan) | Three cun below the middle of the navel, on the front median line |

#### 2.7.1 Acupuncture group

According to the intervention standard of a RCT, the subjects were treated with manual acupuncture and the insertion acupuncture was performed with the Park devices. Firstly, after skin disinfection, it is preferred to prick GB20 (Fengchi) point without retaining the needle. Secondly, the rest manipulation of acupuncture is as follows: ① The scalp needles: GV20 (Baihui), GV29 (Yintang), EX-HN1 (Sishencong); ②The six acupoints of manual: auricular points (bilateral): CO15 (Heart), CO14 (Lung) and TF4 (Shenmen); Body points (bilateral): LI20 (Yingxiang), EXTRA12 (Anmian) and ST36 (Zusanli) are the main points; ③ The three acupoints of sleep (bilateral): (HT7 Shenmen), PC6 (Neiguan) and SP 6 (Sanyinjiao); ④ Siguan points (bilateral): LI4 (Hegu), LR3 (Taichong); ⑤The abdominal four acupoints: bilateral ST25 (Tianshu), CV12 (Zhongwan), CV4 (Guanyuan). The body points were applied with needles of 0.3mm*40mm, and the depth of needling is at 0.5-1 cun. After obtaining Qi, the points were manipulated with the mild reinforcing-reducing method. The auricular was inserted with a 0.25mm*40mm needle, and only straight needling is performed for 0.2-0.3 cun. The needles are retained for 30 min, and the treatment will be given once every other day, 3 times a week. The effect is evaluated at different time points (check in figure 1). The distribution of each acupoint is shown in table 1.

#### 2.7.2 Placebo acupuncture group

Subjects in the placebo acupuncture group will receive non-insertive acupuncture using the sham needle supported by the Park device. This needle has a retractable shaft and a blunt tip, they could not penetrate the skin. We gently placed the sham needle and Park device on the skin. The sham needle is then no longer manipulated in order to minimize any physiological effects. The selected acupoints and needle retention time were the same as for the acupuncture group. Since oblique insertion is required for acupuncture points on the head, the fixed acupuncture body of the Park device could not be used. Therefore, blunt needles are used to stimulate the head points to reduce the amount of stimulation produced by acupuncture. At the end of the treatment, the acupuncturist also used a dry cotton swab to press the acupoints so that patients could feel the withdrawal of ‘real’ needles[13].

### 2.8 Outcomes

#### 2.8.1 Primary outcomes

With the assistance of researchers, the subjects were evaluated by PSQI scale according to their own conditions before treatment, the 4th week of treatment, and 4 weeks after treatment. The score data were collected to analyze insomnia scores changes before and after treatment. PSQI was used to assess sleep disorders through subjective sleep quality, sleep duration, sleep latency, habitual sleep efficiency, sleep disturbance, daytime dysfunction analyze insomnia scores changes during treatment. Each item is scored on a 4-point scale (0-3), and the total score ranges from 0 to 21. The higher the score means better quality of sleep, the lower the sleep quality poorer quality of sleep.

#### 2.8.2 Secondary outcomes

Secondary outcomes mainly included the following:

(1) Chalder-14 Fatigue scale and Epworth Sleepiness Scale were used to evaluate the degree of fatigue after awakening before treatment, at the 4th week of treatment, and 4 weeks after treatment. The score data will be collected to analyze the change of the degree of fatigue after waking up before and after treatment. The Chalder-14 Fatigue Scale consisted of 14 items, which reflect the severity of fatigue from different aspects. The 14 items are divided into two categories, one reflecting physical fatigue and the other reflecting mental fatigue. The highest score of physical fatigue is 8 points, the highest score of mental fatigue is 6 points, and the highest total score is 14 points. The higher the score, the more severe the fatigue. The Epworth Sleepiness Scale is used to assess whether people are excessively sleepy during the day, with higher scores indicating a greater tendency to drowsiness. In particular, 0-8 points is normal, 9-12 points is mildly abnormal, 13-15 points is moderately abnormal, and more than 16 points is severely abnormal.

(2) Emotional disorders rating scales (SAS, SDS)

With the assistance of researchers, the subjects were evaluated by SAS and SDS scale according to their emotional state before treatment, the 4th week of treatment, and the 4 weeks after treatment. The score data were collected to analyze the changes of anxiety and depression scores before and after treatment. The cut-off value of SAS standard score was 50 points, in which 50-59 points were classified as mild anxiety, 60-69 points as moderate anxiety, and more than 70 points as severe anxiety. The cut-off value of SDS is 53 points, of which 53-62 points are mild anxiety, 63-72 points are moderate anxiety, and more than 73 points are severe anxiety. The upper limit of normal for the total crude SDS score is 41, with lower scores indicating better status. The standard score is the integer fraction obtained after multiplying the total crude score by 1.25. In China, the standard score of SDS ≥53 is defined as depressive symptoms.

(3) Evaluation by PSG

All subjects underwent polysomnography (PSG) after 4 weeks of treatment. Philips polysomnography (model: Alice 6) was used to monitor the PSG. The subjects were ready to wash and prepare to wear the electrodes. Positioning and interpretation were performed according to the American Academy of Sleep Medicine Manual for Interpretation of Sleep and Related Events, version 2.6. The international 10-20 system was used to determine the brain area, and the electroophthalmogram and mandibular electromyogram electrodes were determined according to the manual for pre-selection and localization. Then the skin was cleaned with scrub cream, and the electrodes were fixed with conductive cream, and the head was fixed with elastic mesh cap. The electrocardiogram (ECG) leads were fixed with button electrodes, and the chest and abdomen belts were bound in order. A nasal catheter, snore detector, and blood oxygen clip were placed. The operator opened Sleepware G3 data analysis software, after entering the personal information of the subjects, opened the software, opened the record, and began to officially record the data after calibration. Patients were instructed to rest, and the time of lights off and turning on at night was recorded. The data collected by Sleepware G3 were used to analyze the changes of sleep parameters of patients, including sleep efficiency, the proportion of each sleep stage, sleep onset latency, and sleep latency corresponding to each stage.

(4) Assessment of HRV

Dynamic digital electrocardiogram recorder (model: Seer12) and GE MARS software were used to measure HRV data before and after treatment to evaluate the changes of autonomic nerve function. Subjects avoided strenuous activity within 1h before the test, and avoided drinking alcohol, tea, or coffee within 24h before the test. After resting for 10 min, the subjects were placed in a supine position and kept relaxed. Medical alcohol gauze was used to wipe the area where the electrode sheet needed to be attached, and then special sandpaper was used to gently wipe so as to reduce artifacts and impedance. The corresponding electrodes were pasted according to the position where the 12-lead holter electrodes were pasted. After the tape was fixed, the chest lead was fixed with an elastic bandage to reduce the occurrence of artifacts. The instrument was started and the relevant parameter values were recorded. Low-frequency power (LF), high-frequency power (HF), and the ratio of low-frequency to high-frequency power (LF/HF) were analyzed according to the frequency domain analysis. HF reflects parasympathetic function, LF reflects sympathetic and parasympathetic function, and LF/HF reflects sympathetic and parasympathetic balance[14–16].

### 2.9 Safety assessment

Accidents that may occur during treatment, such as dizzy, bent needle, stuck needle, broken needle, subcutaneous hematoma, etc. When adverse events occurred, they were treated according to the operation standard, and their occurrence time, end time, degree, specific treatment measures and outcome were recorded in detail.

### 2.10 Data management and confidentiality

All researchers, including acupuncturists, data managers, data collectors, data entry clerks, and outcome assessors, will receive professional and standard training on data collection and management. All raw data will be clearly and completely preserved in the case report form (CRF), informed consent, and examination report. CRF includes treatment time point, follow-up time point, outcome assessment results, etc. The research staff will fill in the information on time. If any discrepancies are found, they will be corrected based on the original CRF. All paper documents will be kept in a locked filing cabinet, and electronic documents will be kept on a special computer with password protection. Only the corresponding researcher has access to these documents. After publication, all trial-related data will be archived for at least 5 years for use and review.

### 2.11 Quality control

A standard acupuncture procedure needs to be established for the identification, enrollment, and treatment of participants. All practicing physicians are involved in treatment must be trained to ensure consistency throughout the study. A data safety monitoring plan was in place, and all adverse events were recorded, managed, and followed until they resolved. In addition, all adverse events were reported to both the ethics committee.

### 2.12 Sample size

PASS15.0 software was used to estimate the sample size. This study was a completely randomized controlled trial. The PSQI of the subjects was the main outcome index observed. By referring to several literatures, it was predicted that after treatment, the PSQI value of the study subjects was 10.0 in the treatment group and 7.5 in the control group, with a bilateral α = 0.05, a power of 90%. The proportion of the patients in the two groups was set as follows (the acupuncture group: the placebo acupuncture group = 1:1). PASS15.0 software was used to calculate the sample size of 32 cases in the acupuncture group and 32 cases in the placebo acupuncture group. Cases lost in this study should be taken into consideration, the number of patients in the two groups was set to be 37 on the basis of 15% loss of subjects.

### 2.13 Statistical analysis

The data in this study were analyzed using SPSS16.0 software, and the measurement data were expressed as mean ± standard deviation (x±S): Independent sample t test was used for normal distribution and homogeneity of variance, corrected t test was used for normal distribution but uneven variance, and Wilcoxon Mann-Whitney method was used for rank sum test for non-normal distribution. The paired sample t test was used to compare the measurement data within the group. Count data were expressed as frequency and rate, and non-rank count data were analyzed by χ^2^ test. In this study, *P<0.*05 was considered to be statistically significant.

## 3. Discussion

According to traditional Chinese medicine theory, the etiology of PI is closely related to mental disorder, negative emotions such as anxiety and depression[17,18]. The manual acupuncture can regulate brain function and adjust mood. So far, there is no standardized clinical research to explore the therapeutic effect and mechanism of manual acupuncture on the PI. This study is designed as a prospective, single-center, double-blind randomized controlled trial. By using placebo acupuncture as a control, this study aims to further clarify the clinical superiority of the manual acupuncture in treating the PI, and at the same time explain the mechanism of the manual acupuncture. It provides a scientific theoretical basis for the clinical promotion of the manual acupuncture.

The PI is defined as difficulty in initiating or maintaining sleep with associated impaired daytime energy or work capacity despite adequate sleep opportunities and a suitable sleep environment is available. With the accelerated pace of life and increasing social stress, the prevalence of PI is gradually increasing. The theory of regulating the mind refers to the theory of using relevant acupuncture points for regulating the autonomic nerve to regulate the functions of the meridians and zangfu, thereby improving insomnia[19]. Moreover, modern medicine indicates that acupuncture can relieve PI symptoms by regulating the excitability of the autonomic nerves[20,21].

### Selected acupoints

The manual acupuncture focuses on the idea of “regulating the mind first”, advocating the treatment of insomnia with the “mind-regulating theory” as the core, combining with vagus nerve stimulation through auricular points, and finally matching points according to the syndrome, which can quickly improve the sleep quality of PI patients[22–24]. It is emphasized in this theory that the key to regulating the mind is to select the acupoints on the head, namely GV20, GV29, GB20, to regulate the brain function directly[25,26]. Then open the Siguan points: bilateral LI4, LR3, to regulate the nutrient and superficial defensive system; take Six points for calming the nerves, that is bilateral auricular points: CO15, CO14, and TF4. Auricular point can directly stimulate vagus nerve afferent fibers can generate the potential which may originate from the brain stem vagus dorsal nucleus and nucleus multipolar neurons in the brain. The signal can contact with the hypothalamus, amygdala, and the cholinergic node fiber, control secretion melatonin, and finally improve sleep quality[21]. Moreover, auricular point can activate the release of endogenous opioids, such as enkephalin, β-endorphin, and endomorphin, which regulate the processing of pain signals and calm the nerve[27]. Body points have been proved to have different functions, like LI20 can relieve anxiety, EXTRA 1 point can make it easier to fall asleep, ST36 regulate Qi of stomach; take four abdominal points: ST25 (bilateral), CV12, CV4, modulate state of nerve, and regulate Qi and blood[28]. Finally, through matching additional points according to the syndrome, we’ll take ST4 if the gastric fire disturbs the heart, and take ST40, SP9 to against turbid phlegm[29].

### Innovativeness of this trial

We noticed the close connection between sleep quality and mood, then both sleep quality and changes in mood disorders were detected. We observed that scales were often used as the main evaluation method in previous studies of acupuncture treatment on sleep disorders[30,31]. In this study, in addition to the conventional sleep quality and emotional disorder evaluation scales, we also use PSG to observe the changes in sleep quality objectively, and apply HRV to observe the changes in the autonomic nervous function of the PI patient, so as to objectively estimate the sleep quality and emotional changes of PI patients treated with the manual acupuncture. By combining subjective and objective data, we comprehensively judge the therapeutic effect of the manual acupuncture on sleep and emotion of PI patients and analyze its potential mechanism[16].

### Design of placebo needle

It’s been proved that Park device has minimal stimulation to the human body, has no specific therapeutic effect on the disease, which has a strong ability to mask patients. What needs to be emphasized is that the Park device mainly used in the body and body relatively flat parts, and the blunt needles are used at head points to reduce acupuncture effects. At the same time covers the patient’s visual field, reducing the patient’s suspicion of the needling process. In this experiment, both of them are combined to achieve blinding under the premise of ensuring safety.

However, this RCT will inevitably have some limitations. Firstly, PI is a chronic disease and therefore requires long-term and single center observation, while this trial merely focuses on short-term efficacy. In addition, the sample size of this trial was only 37 subjects per group, which may have limited the reality bias and generality of our study hypotheses. Therefore, we should establish a long-term and multi-center study with a larger sample size for verification, and exploration in the future.

## Data Availability

No datasets were generated or analysed during the current study. All relevant data from this study will be made available upon study completion.

## List of abbreviations

PI: primary insomnia
CBTi: cognitive behavioral therapy for sleep
PSQI: Pittsburgh sleep Quality Index
PSG: polysomnography
HRV: heart rate variability
ICSD-3: International Classification of Sleep Disorders third edition
CRF: case report form.

## Authors’ contributions

All authors contributed significantly to the reported work in all areas. The original idea was conceived by YL. YL, KZ drafted the manuscript for this protocol. BW, QZ, DH, WC and WW participated in the design of the study and the setting of the inclusion and exclusion criteria. LY, KZ will be in charge of data acquisition, analysis and interpretation. XL, CZ will review all the work. All authors have read and approved the publication of the protocol and consent to submit to the journal.

## Availability of data and materials

No data has been generated at this time. Data from RCTs will be obtained from the corresponding authors upon reasonable request.

## Acknowledgments

All the subjects who participate in this trial will be appreciated.

## Supporting information

**S1 Fig.Flowchart of trail procedures.** This is the S1 Fig legend.

**S2 Fig.Park device operation diagram.** This is the S2 Fig legend.

